# Alterations in resting-state activity and functional connectivity in children with 22q11.2 deletion syndrome

**DOI:** 10.1101/2021.09.14.21263530

**Authors:** Joanne L. Doherty, Adam C. Cunningham, Samuel J.R.A. Chawner, Hayley M. Moss, Diana C. Dima, David E. J. Linden, Michael J. Owen, Marianne B.M. van den Bree, Krish D. Singh

## Abstract

**Background:** While genetic risk factors for psychiatric and neurodevelopmental disorders have been identified, the neurobiological route from genetic risk to neuropsychiatric outcome remains unclear. 22q11.2 deletion syndrome (22q11.2DS) is a copy number variant (CNV) syndrome associated with high rates of neurodevelopmental and psychiatric disorders including autism spectrum disorder (ASD), attention deficit hyperactivity disorder (ADHD) and schizophrenia. Alterations in neural integration and cortical connectivity have been linked to the spectrum of neuropsychiatric disorders seen in 22q11.2DS and may be a mechanism by which the CNV acts to increase risk. Despite this, few studies have investigated electrophysiological activity and connectivity in this high-risk group.

**Methods:** Magnetoencephalography (MEG) was used to investigate resting-state cortical oscillatory patterns in 34 children with 22q11.2DS and 25 controls aged 10-17 years old. Oscillatory activity and functional connectivity across six frequency bands were compared between groups. Regression analyses were used to explore the relationships between these measures, IQ and neurodevelopmental symptoms.

**Results:** Children with 22q11.2DS had atypical oscillatory activity and functional connectivity across multiple frequency bands. In the 22q11.2DS group, low frequency (alpha band) activity was positively associated with cognitive ability, while connectivity was negatively associated with ASD and ADHD symptoms. Frontal high frequency (gamma band) activity and connectivity were positively associated with ASD and ADHD symptoms respectively, while posterior gamma activity was negatively associated with ASD symptoms.

**Conclusions:** These findings highlight that haploinsufficiency at the 22q11.2 locus alters both local and long-range cortical circuitry, which could be a mechanism underlying neurodevelopmental vulnerability in this high risk group.

## Introduction

Copy number variants (CNVs) are submicroscopic alterations (such as deletions or duplications) of segments of chromosomes, which constitute a major source of variation between individuals. CNVs can result in chromosomes having too many or too few dosage-sensitive genes. These changes may be advantageous, indeed CNVs are thought to have been important in human evolution (1); however, they may also have a negative impact on human development (2).

22q11.2 deletion syndrome (22q11.2DS), also known as velocardiofacial syndrome (VCFS) and Di George syndrome, is one of the most common copy number deletion syndromes, affecting at least 1 in 4000 live births (3–5). It results from a 1.5-3Mb deletion at region q11.2 on chromosome 22. As with many CNV syndromes, the 22q11.2DS phenotype is extremely variable, involving multiple organ systems (6). The mechanisms underlying the pleiotropy and phenotypic variability in 22q11.2DS are, as yet, not well understood. Neurodevelopmental, behavioral, emotional, cognitive and psychiatric problems are very common in 22q11.2DS (7–10). The 22q11.2 International Consortium on Brain and Behavior study of over 1400 individuals with 22q11.2DS found that 21% of children with the deletion have ASD, 32% have ADHD and up to 30% develop schizophrenia in adulthood (11).

The 22q11.2 region contains approximately 60 genes, but as yet no gene or combination of genes has been found to be either necessary or sufficient for the neurodevelopmental phenotypes observed in the deletion syndrome. It has been proposed that 22q11.2DS and other neurodevelopmental CNVs increase risk of psychopathology by affecting neuronal integration with downstream effects on local and long-range cortical networks (12). There is mounting evidence from animal models for abnormal neuronal structure, function and migration (13–15), as well as abnormal cortical network activity in 22q11.2DS (16,17). In humans, postmortem studies have also found abnormal neuronal morphology and migration in 22q11.2DS (13–15,18,19), while structural magnetic resonance imaging (MRI) studies report reductions in cortical surface area (20), subcortical volumes (21) and white matter diffusivity (22). Furthermore, resting-state functional MRI (fMRI) studies have found alterations across several brain networks including the default-mode, sensorimotor, visuospatial, self-referential and visual networks (23–27). Alterations in neurotransmitter concentrations have also been reported in adults with 22q11.2DS and these have been associated with psychotic symptoms (28,29).

Synchronous oscillatory activity in the cortex reflects neuronal integration and is thought to facilitate communication within and between brain regions (30,31). It can be measured non-invasively in humans using EEG and MEG. Atypical oscillatory patterns have been observed in idiopathic ASD (32–34), ADHD (35–37) and schizophrenia (38–40). Despite the proposed neuronal integration deficits in 22q11.2DS and other neurodevelopmental CNVs, there have been relatively few studies of cortical oscillations in these high-risk groups. In a recent resting-state MEG study in adult CNV carriers, we found common patterns of oscillatory dysconnectivity across nine different neurodevelopmental CNV loci, including 22q11.2DS, suggesting a putative mechanism by which neurodevelopmental CNVs could increase risk of psychopathology (41). Due to the relatively small numbers of participants with each CNV, it was not possible to explore the associations between oscillatory activity and neurodevelopmental symptoms or psychopathology. To our knowledge, there have been no previous studies of resting-state cortical oscillations in children with 22q11.2DS. In the present study, we focused exclusively on children with 22q11.2DS, to investigate the impact of this genetic risk factor on oscillatory activity in the developing brain and to explore associations with cognitive function and neurodevelopmental symptoms.

## Methods and Materials

### Participants

Participants were recruited through National Health Service (NHS) genetics clinics and patient support groups in the United Kingdom. 39 children aged between 10-17 years old with a diagnosis of 22q11.2DS and 26 siblings of children with neurodevelopmental CNVs (controls) were recruited to the study. Written consent was obtained in accordance with The Code of Ethics of the World Medical Association (Declaration of Helsinki) and all procedures were approved by the South East Wales NHS Research Ethics Committee. Participants over the age of 16 years old with capacity to consent gave written informed consent to participate in the study. Children under the age of 16 years old or those over the age of 16 who lacked capacity to consent for themselves gave written and/or verbal assent to take part and their parents or carers provided consent for their participation. Participants were excluded from the study if they had photosensitive epilepsy, orthodontic braces or metallic implants/prostheses in the upper half of the body. Participants taking psychotropic medication were excluded from the analyses (n=1).

### Genotyping

CNV status was confirmed for all participants. Affected children were excluded from taking part in the study if a 22q11.2 deletion was not confirmed either by in-house testing at the MRC Centre for Neuropsychiatric Genetics and Genomics at Cardiff University or by clinical genetics report. All control participants had in-house microarray testing and were excluded from the analysis if they carried a neurodevelopmental CNV as defined in (42) (n=1). Genotyping was carried out using the Illumina HumanCoreExome whole genome SNP array, which contains 27000 additional genetic variants at loci previously linked to neurodevelopmental disorders, including CNVs.

### Cognition and psychopathology

Psychiatric and cognitive data were collected from participating children and their primary caregiver by trained research psychologists, supervised by child and adolescent psychiatrists. Psychiatric interviews were conducted with the primary carer using the Child and Adolescent Psychiatric Assessment [CAPA; (43)] to derive DSM-5 (44) ADHD diagnoses and symptom counts. The Social Communication Questionnaire [SCQ; (45)] was completed by the primary carer and used to assess ASD symptoms. Children’s full-scale IQ scores were obtained using the Wechsler Abbreviated Scale of Intelligence [WASI; (46)]. Further details about the clinical and cognitive phenotyping protocol have previously been reported (9).

### Data acquisition

Five minute whole-head MEG recordings were acquired at a 1200Hz sample rate using a 275- channel CTF radial gradiometer system. An additional 29 reference channels were recorded for noise cancellation purposes. Primary sensors were analysed as synthetic third-order gradiometers (47). Participants were seated upright in the MEG system during the recordings and were instructed to keep their eyes open and to attend to a red fixation point presented on a mean luminance background using a Mitsubishi Diamond Pro 2070 monitor (100Hz frame rate) or PROPixx LCD projector (120Hz frame rate). Electromagnetic coils were placed at three fiducial locations (bilateral preauricular regions and nasion) and their position relative to the MEG sensors was localised at the beginning and end of each recording. Relative head position at the beginning and end of the recording was used as a proxy measure of participant head motion.

After the recordings, MEG data were down-sampled to 600Hz, band-pass filtered at 1-150Hz and segmented into 2s epochs, generating 150 trials per participant. Data were then visually inspected for gross artefacts such as motion, muscular contraction and eye movements. Trials containing such artefacts were removed from the dataset and excluded from further analysis. Participants with head motion greater than 30mm or with greater than half of their trials containing artefacts were excluded from further analysis (n=4).

After quality control, data from 34 children with 22q11.2DS (17 female) and 25 controls (10 female) remained and were included in the subsequent analyses. There were no significant between-group differences in head motion (median=4.65mm (IQR=5.4mm) in children with 22q11.2DS, median=2.9mm (IQR=4.6mm) in controls, p=0.61) or the number of trials after artefact rejection (median=131 (IQR=34) in children with 22q11.2DS, median = 133 (IQR=29) in controls, p=0.22).

Where possible, T1-weighted MRI structural images were acquired either with a 3D fast spoiled gradient echo (FSPGR) sequence on a General Electric HDx MRI system (TR=7.8ms, TE=3.0ms, voxel size = 1mm isotropic) or a 3D magnetization prepared rapid acquisition gradient echo (MP-RAGE) sequence on a Siemens Prisma MAGNETOM MRI system (TR=2.3ms, TE=3.06ms voxel size = 1mm isotropic). Co-registration was performed by manually labelling the fiducial points on each participant’s MRI using the software package MRIViewer. T1 MRI data were not available for 15 participants due to MRI contraindications or poor data quality (e.g. due to participant head motion). For these participants, an appropriate MRI scan was selected from the available data by comparing the relative distances between the fiducial points for each participant and matching these with another participant’s dataset. The resulting co-registrations were visually inspected and a good quality match was achieved in all cases.

### MEG Analysis: Activity and connectivity estimation

The analysis pipeline is described in detail in supplementary material and summarized in Figure 1. Source reconstruction was performed using a linearly-constrained minimum variance (LCMV) beamformer in FieldTrip (version 20161011, www.fieldtriptoolbox.org, (48). Beamforming was performed in each of six distinct frequency bands using conventional definitions: delta (1-4 Hz), theta (4-8Hz), alpha (8-13Hz), beta (13-30Hz), low gamma (40- 60Hz) and high gamma (60-90Hz). Beamformer weights were used to derive an estimated activity time series at each voxel on a 6mm grid and for each trial. These trial time series were concatenated to form a single time series for each grid voxel and then taken forward for both activity and connectivity analyses.

**Figure 1.**
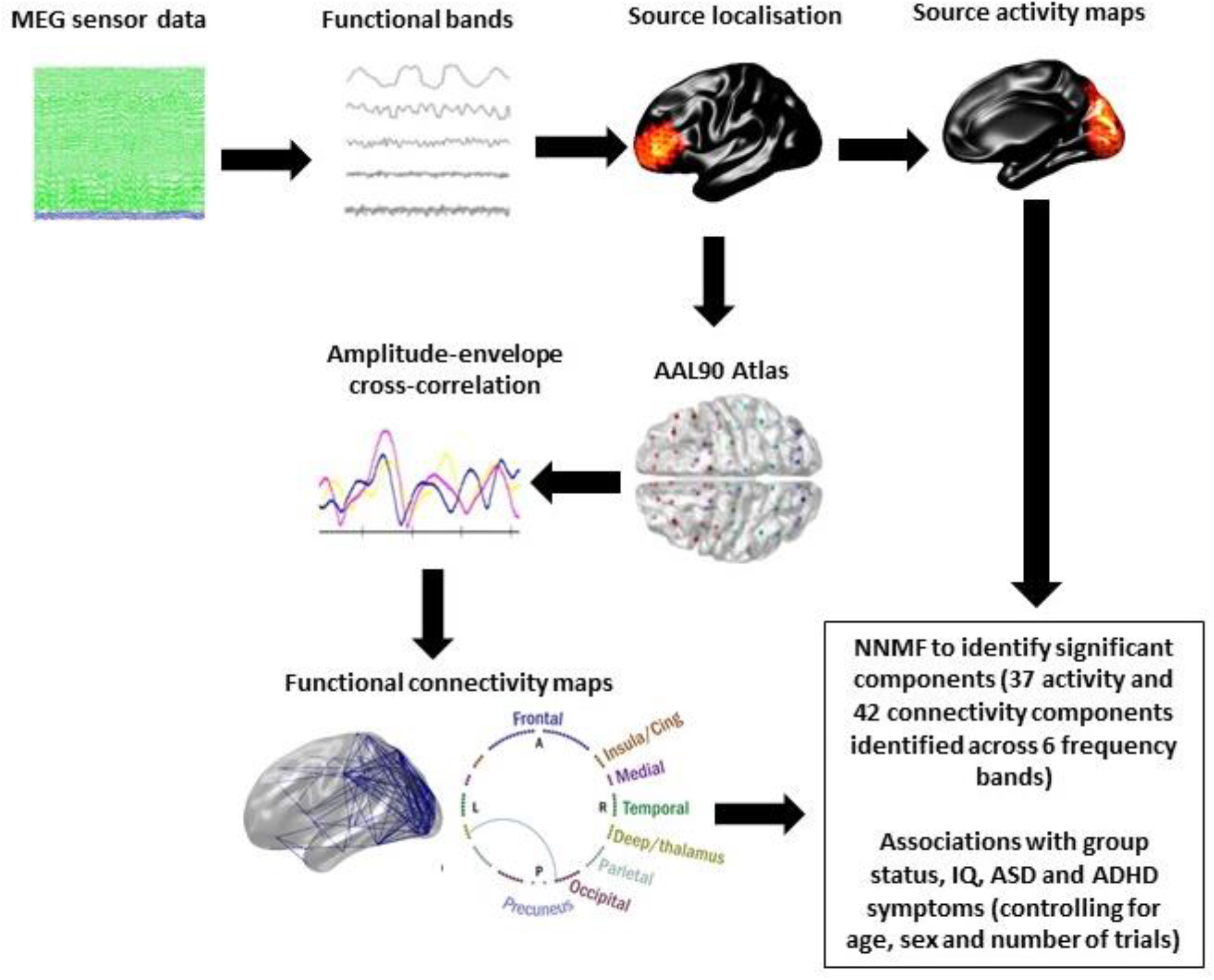
Overview of the MEG analysis pipeline. Sensor level MEG data were filtered into the six frequency bands of interest. Source localization was performed for each participant and each frequency band using a linearly-constrained minimum variance (LCMV) beamformer. Source activity was estimated for each participant and frequency band by calculating the temporal coefficient of variation (CoV) of the Hilbert envelope. For functional connectivity estimates, data were first down-sampled to the 90 regions of the Automated Anatomical Labelling (AAL90) atlas. The temporal activity of each of these 90 sources was then orthogonalized and the Hilbert envelopes of each AAL90 region were extracted. These amplitude envelopes were then down-sampled to a temporal resolution of 1s and a median spike removal filter was applied. Pairwise correlations were calculated between the 90 Hilbert envelopes, for each frequency band and each participant. Non-negative matrix factorization was performed to identify the principal components for each of the six frequency bands of interest. This reduced the data to 79 components (37 activity components and 42 connectivity components) that were taken forward for further analysis. Relationships between the components, group status, IQ and neurodevelopmental symptoms were explored using regression models with age, sex and number of MEG trials included in the models as covariates.

For each of the reconstructed grid positions, a measure of neural activity was derived in each frequency band by first computing the amplitude envelope of the virtual-sensor time series using the absolute value of the analytic function transform of the raw time series (using Matlab’s *hilbert* function) and then converting this to a single activity measure that summarizes how variable this envelope is over the entire resting-state run. To do this we calculated the coefficient-of-variation of the envelope. This normalized measure has the advantage of correcting for the known biases introduced by the sensitivity of beamformer weights to variations in the signal to noise ratio (SNR) of the data (48,49). The end result is a 6mm isotropic activity map for each participant and each frequency band.

Functional connectivity was computed using the amplitude-envelope correlation (AEC) metric. This metric has previously been shown to be both robust and repeatable (50). The analysis pipeline has previously been described (48). First, spatial down-sampling to the 90 regions of the Automated Anatomical Labelling (AAL90) atlas was performed (51). The temporal activity of each of these 90 sources was then orthogonalized with respect to each other in order to suppress any zero-time-lag correlation due to signal leakage (52). Next, the amplitude (Hilbert) envelopes of each AAL90 region were extracted using the absolute of the (complex) analytical signal derived by the *hilbert* function in MATLAB. These amplitude envelopes were then down-sampled to a temporal resolution of 1s in order to study connectivity mediated by slow amplitude envelope changes (53). To obtain connectivity matrices, pairwise correlations were calculated between the 90 Hilbert envelopes, yielding 4005 unique correlations for each frequency band for each participant. Each of these correlation coefficients was then transformed to a variance-normalized Fisher z-statistic. This made the correlations suitable for further statistical analysis and corrected for the varying length of the final time series for each participant.

### MEG analysis: Activity and connectivity component estimation

At the end of the above analysis procedures, each participant had 6 activity maps and 6 connectivity matrices (one for each frequency band). Each activity map had 5061 voxels and each connectivity map had 4005 unique connection values. In order to reduce the dimensionality of these features before statistical analyses, we used a data-driven analysis of the principal components using non-negative matrix factorization (NNMF, Matlab: *nnmf*). Recently, non-negative matrix factorization has been successfully used to show cohort differences in a MEG study of schizophrenia (54) and in comparing structural and functional connectivity components in healthy individuals (55). We iteratively increased the number of components and tested what proportion of our cohort had non-zero values. We required each component to be represented in at least 50% of our participants and for the mean number of participants represented to be at least 70%. For each of the final components identified, we projected each individual’s data on to these networks to get a single component ‘strength’ for each person. For each of the 12 metrics we have in each person (6 activity and 6 connectivity), we performed NNMF separately. As shown in figures 1,2 and 7, each participant’s combined activity and connectivity profile, across all 6 bands, was effectively summarised by just 79 values. It is these values that were taken forward for statistical analysis.

### Statistical analyses of NNMF derived component scores

Each of the component weightings described above was used in an analysis to determine whether their magnitude was predicted by a set of exploratory variables consisting of group status (22q11DS or control), IQ, SCQ score and ADHD symptoms. Due to differing IQ distributions in the two groups, associations with IQ were explored in each group separately. This analysis was done by a set of univariate robust general linear modelling tests, using

Matlab’s *fitlm* function. In each linear-model fit age, sex and number of MEG trials were included in the models as covariates. For linear models exploring the associations with SCQ scores and ADHD symptoms, IQ was included as an additional covariate. For each test, we assessed the significance of the principal variable in explaining variance in the residuals. Effect-sizes for the principal variable of interest were calculated using standardised beta parameters and assessed for significance using p-values and 95% confidence intervals. Bonferroni correction was applied for the number of components tested within each type (activity/connectivity) and frequency band. To control for relatedness between sibling pairs in the sample, an additional sensitivity analysis using linear mixed modelling was run in R Studio (Version 1.1.383 for Mac) for each of the significant components using the package lmerTest. In these analyses, component weighting was the variable of interest with group status, age, sex and number of MEG trials as fixed effects and family identification numbers as random effects.

## Results

### Participant characteristics

Table 1 shows demographic, cognitive and clinical characteristics of the 34 children with 22q11.2DS and 25 controls who were included in the analyses. Children with 22q11.2DS had full-scale IQ scores that were shifted approximately 30 points to the left of typically developing controls. 29.4% of children with 22q11.2DS scored above 15/40 in the SCQ, which is a commonly used screening cut-off for a likely ASD diagnosis (57). 17.6% of children with 22q11.2DS met DSM-5 criteria for ADHD on the basis of symptoms reported in the CAPA. No controls met criteria for either ASD or ADHD diagnoses.

**Table 1.** Participant characteristics

|  | **22q11.2DS** | **Controls** | **Test statistic (t/$\chi^2$)** | **P** |
| --- | --- | --- | --- | --- |
| **Age, mean in years (range, SD)** | 13.5<br>(10.5 -17.5, 1.9) | 14.4<br>(10.5 -17.9, 1.8) | -1.83 | 0.07 |
| **Gender, % female (n)** | 50<br>(17) | 56<br>(14) | 0.04 | 0.85 |
| **Handedness, % right (n)** | 76.5<br>(26) | 76.0<br>(19) | <0.01 | 1.00 |
| **FSIQ score, mean (range, SD)** | 74.4<br>(53-117, 12.9) | 107.4<br>(86-139, 11.0) | 10.45 | $1.3 \times 10^{-14}$ |
| **SCQ score, mean (range, SD)** | 11.0<br>(3-25, 6.4) | 2.4<br>(0-9, 3.1) | 6.76 | $1.32 \times 10^{-8}$ |
| **Likely ASD diagnosis, % (n)** | 29.4<br>(10) | 0<br>(0) | * | * |
| **ADHD symptom score, mean (range, SD)** | 4.1<br>(0-13, 4.2) | 0.3<br>(0-3, 0.7) | 5.33 | $5.42 \times 10^{-6}$ |
| **Likely ADHD diagnosis, % (n)** | 17.6<br>(6) | 0<br>(0) | * | * |

\*Unable to calculate test statistic ( $\chi^2$ ) or p value due to zero cell count in controls

### Source activity

Figure 2 shows the significant components identified in the analysis of source activity for the between-group comparisons and the associations with IQ and neurodevelopmental symptoms. For each of the six frequency bands investigated, several components were identified. The components showing significant associations with group status, IQ (shown separately for children with 22q11.2DS and controls) and ADHD/ASD symptoms in children with 22q11.2DS are highlighted, with red indicating a positive association and blue a negative association. The standardised beta value for the regressions was used as a marker of effect size.

**Figure 2.**
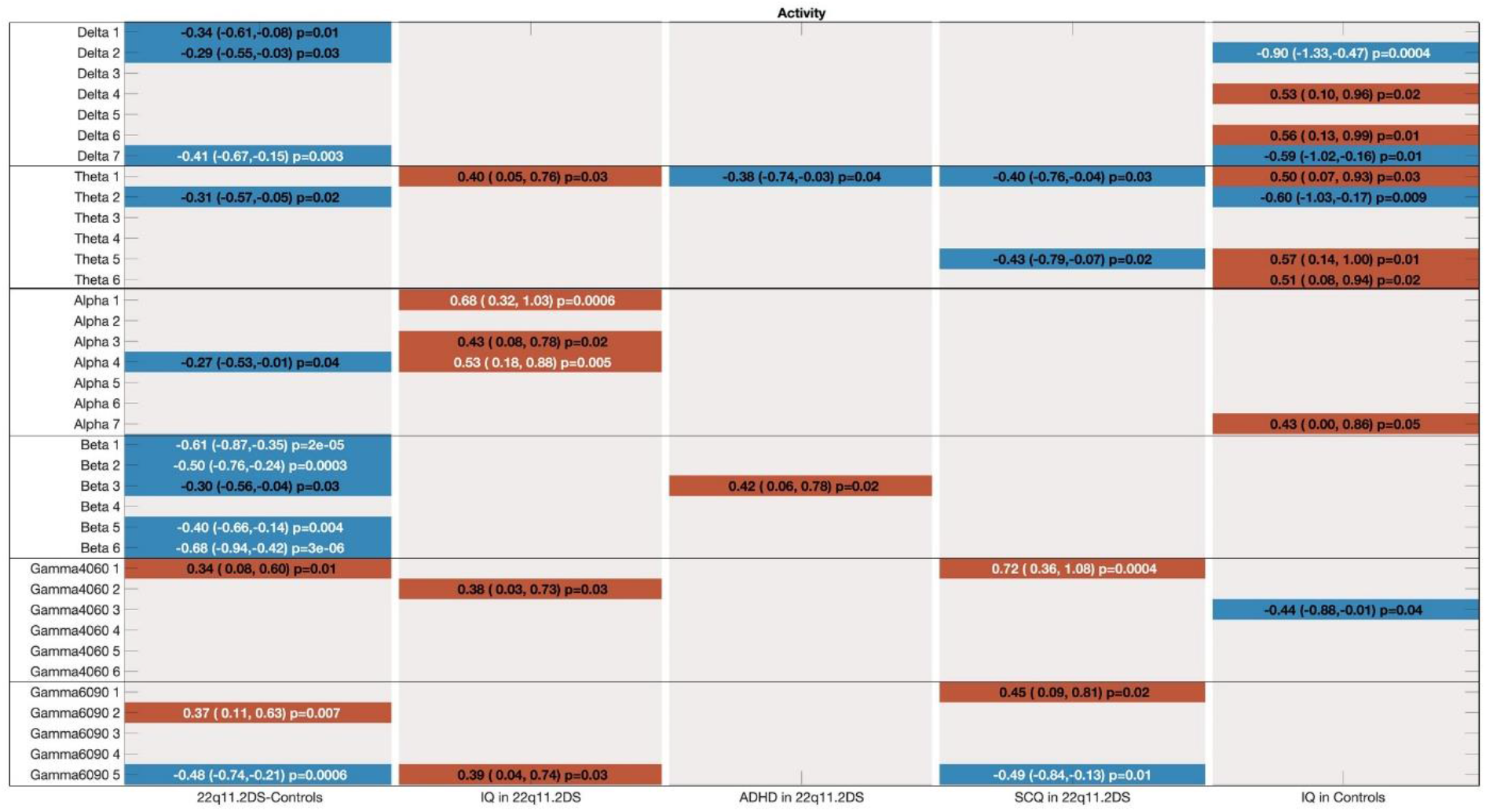
Significant associations between group status, IQ, neurodevelopmental symptoms and source activity for each of the six frequency bands of interest. Components (p<0.05) with positive associations are shown in red, while negative associations are shown in blue. The associations surviving Bonferroni correction are highlighted in white text. Associations are shown by standardised beta values, confidence intervals and p values. Age, sex and number of MEG trials were included in each model as covariates, IQ was included as an additional covariate in models testing associations with SCQ and ADHD scores.

As can be seen in figure 2, significant between-group differences in source activity were evident in the beta, delta and high gamma bands. These differences remained significant when sibling relatedness was taken into account. In the beta band, four components were identified as having significantly reduced oscillatory activity in children with 22q11.2DS compared to controls, with moderate to strong effect sizes of between -0.40 to -0.68. These components were located in occipital and temporoparietal regions (see figure 3).

**Figure 3.**
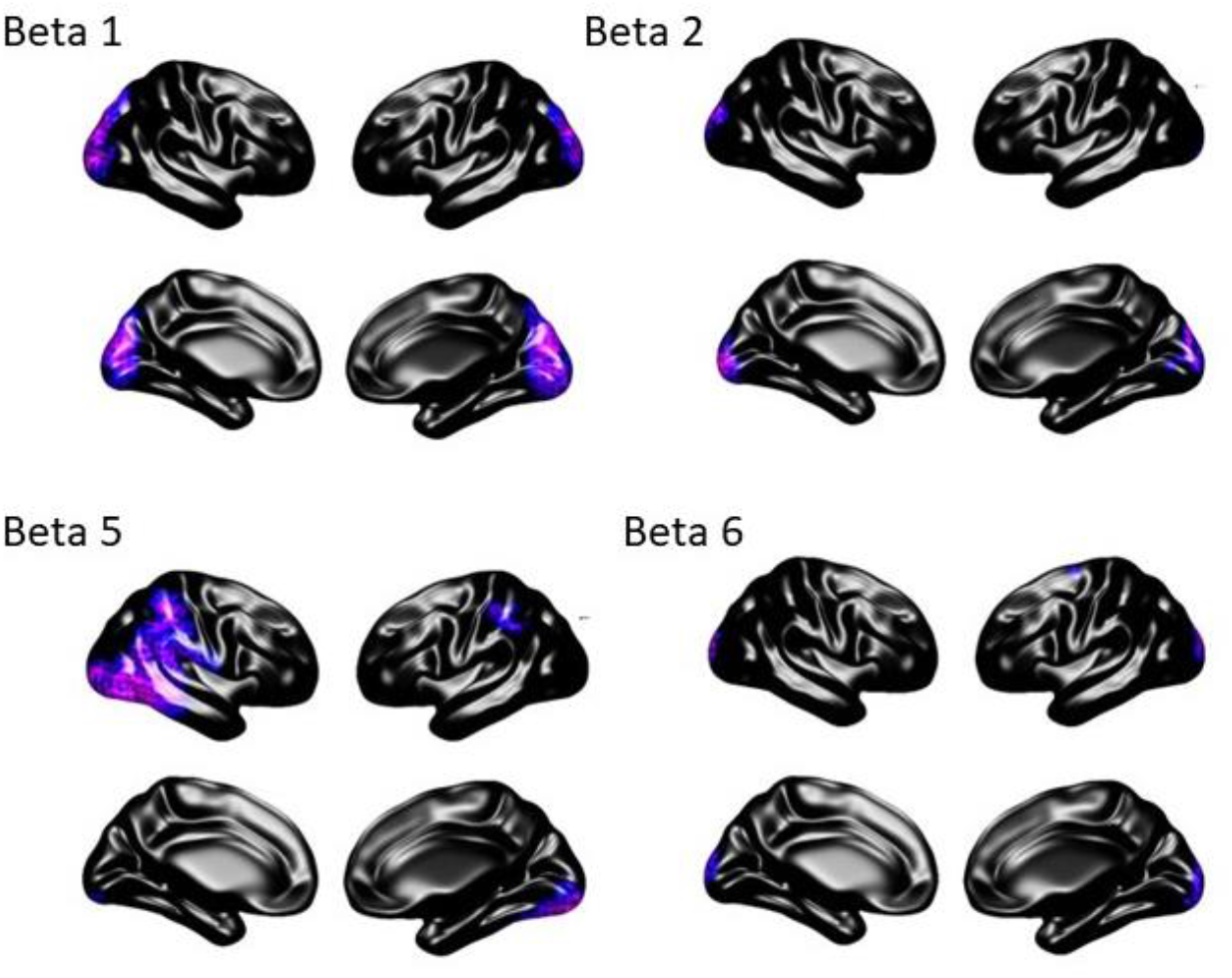
Locations of the four components showing significant between-group differences in beta band activity between children with 22q11.2DS and controls. Purple indicates reduced activity and orange indicates increased activity – all components here showed reductions. Effect sizes: Beta 1 = -0.61 (p=0.000023), Beta 2 = -0.50 (p=0.000332), Beta 5 = -0.40 (p=0.003757) and Beta 6 = -0.68 (P=0.000003). Age, sex and number of MEG trials are included in the models as covariates.

Delta band activity in the medial frontal cortex was reduced in children with 22q11.2DS. In the high gamma band, there were two components with moderate effect sizes. High gamma oscillatory activity was significantly lower in the occipital lobes but significantly higher in the frontal lobes of children with 22q11.2DS. There were no statistically significant differences in oscillatory activity in other frequency bands.

**Figure 4.**
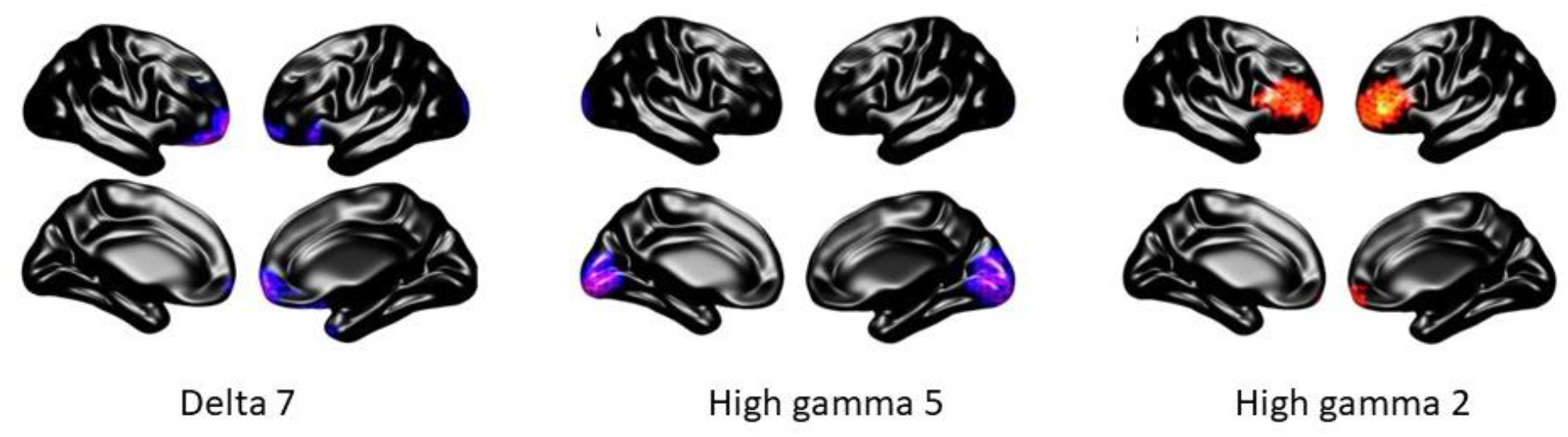
Locations of the delta and high gamma components showing significant between-group differences in source activity between children with 22q11.2DS and controls. Purple indicates reduced activity and orange indicates increased activity. Effect sizes: Delta 7 = -0.41 (p=0.0027), High gamma 5 = -0.48 (p=0.000615) and high gamma 2 = 0.37 (p=0.006592). Age, sex and number of MEG trials are included in the models as covariates.

### Associations between source activity, IQ and neurodevelopmental symptoms

In children with 22q11.2DS, IQ was positively associated with two alpha band activity components located in the occipital and parietal lobes, while in controls, IQ was negatively associated with delta band activity in frontotemporal regions.

**Figure 5.**
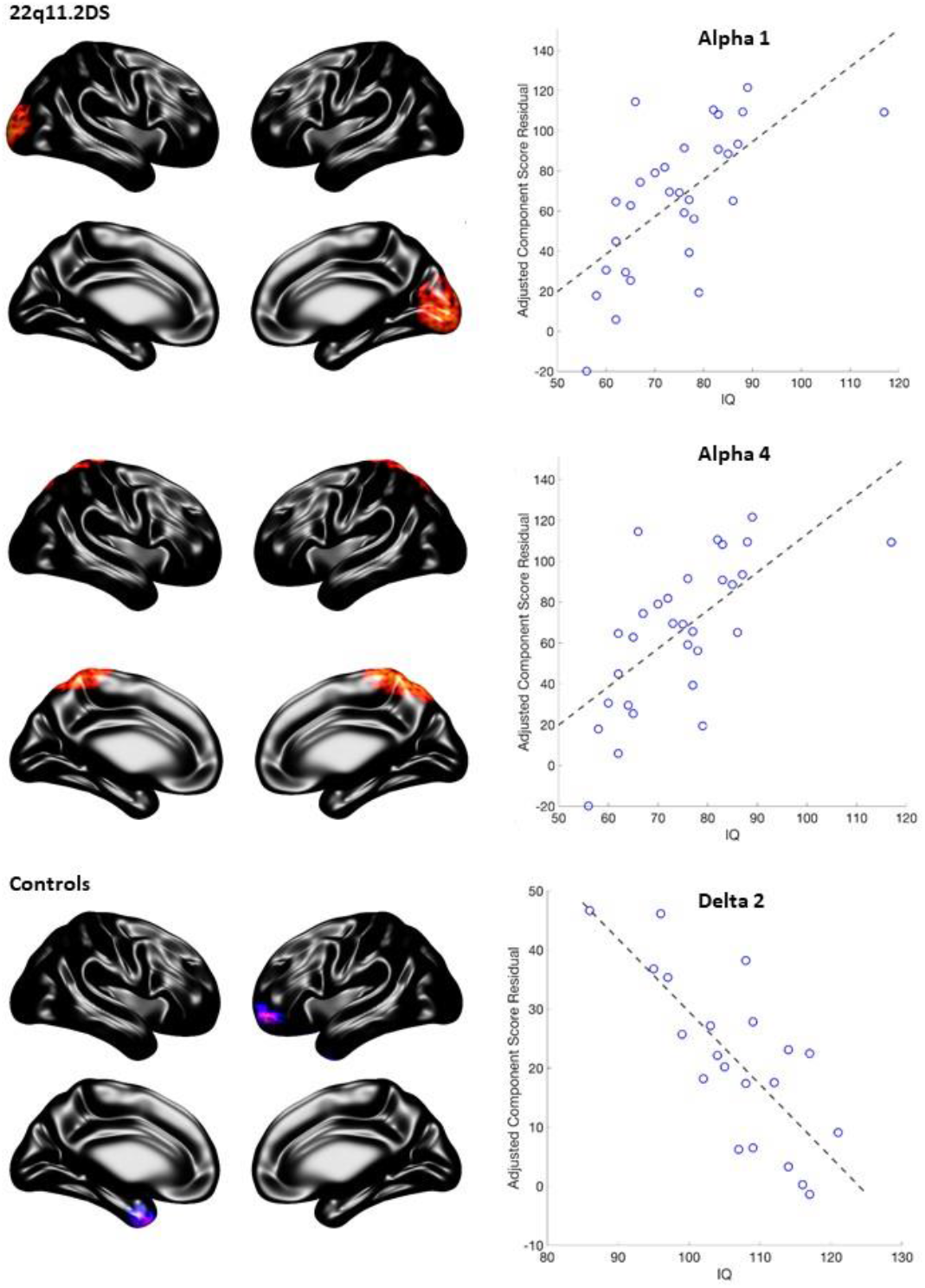
Relationships between IQ and source activity in children with 22q11.2DS and controls, controlling for age, sex and number of MEG trials. Purple indicates reduced activity and orange indicates increased activity. Effect sizes: Alpha 1 (22q11.2DS) = 0.68 (p=0.00058), Alpha 4 (22q11.2DS) = 0.53 (p=0.0046) and delta 2 (controls) = 0.88 (p=0.00044).

There was a significant relationship between gamma oscillatory activity and ASD symptoms in children with 22q11.2DS, with the severity of social communication difficulties being positively associated with frontal gamma activity and negatively associated with occipital gamma activity. There were no significant associations between source activity and ADHD symptoms that survived correction for multiple comparisons.

**Figure 6.**
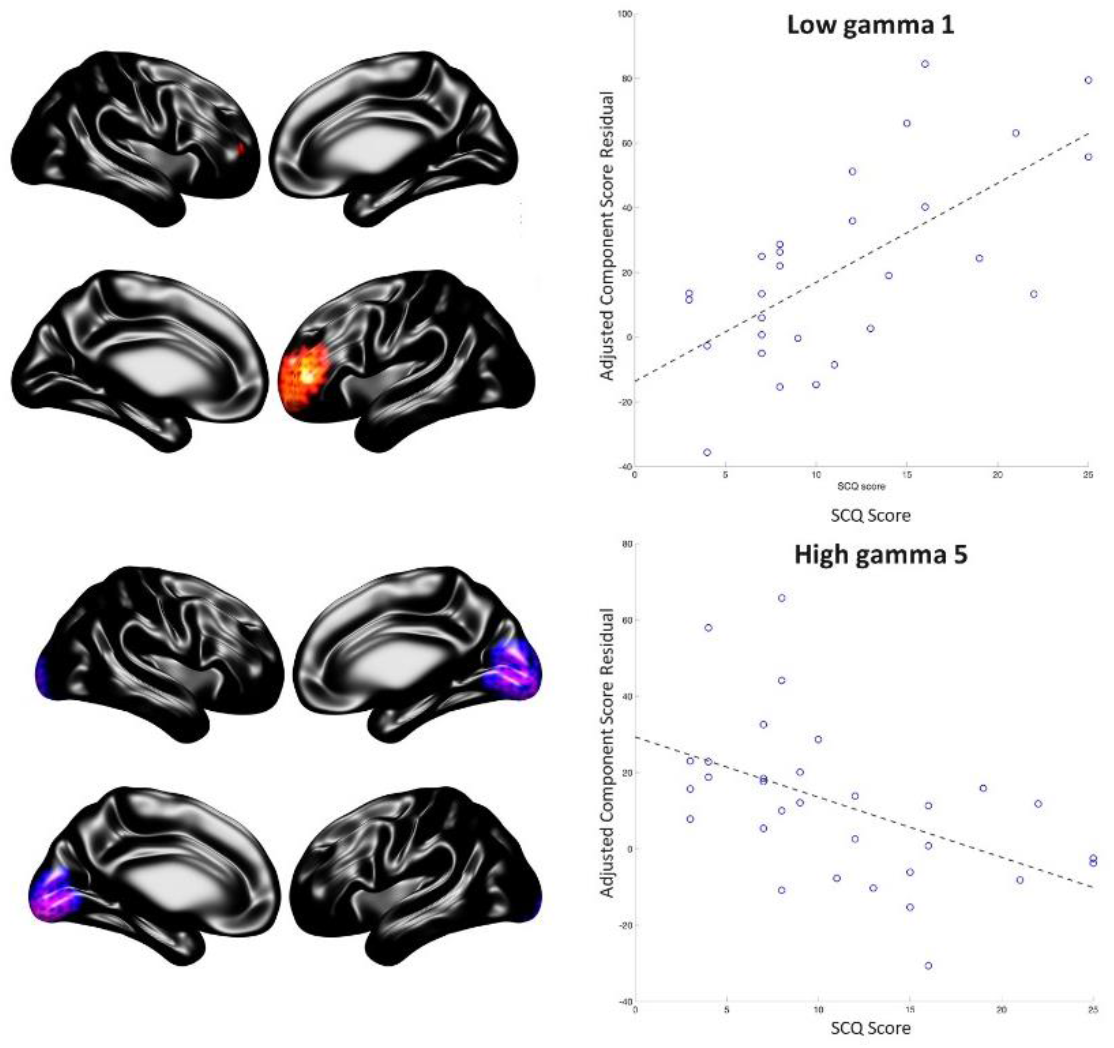
Relationship between SCQ score and gamma activity, controlling for age, sex, IQ and number of MEG trials. Purple indicates reduced activity and orange indicates increased activity. Effect sizes: Low gamma 1 = 0.72 (p=0.0004), high gamma 5 = -0.49 (p=0.01).

### Functional connectivity

Figure 7 shows the significant components identified in the analysis of functional connectivity for the between-group comparisons and the associations with IQ and neurodevelopmental symptoms. As for the analysis of source activity, those components showing significant associations with group status, IQ (shown separately for children with 22q11.2DS and controls), ADHD and ASD symptoms in children with 22q11.2DS are shown in the figure, with red indicating a positive association and blue, a negative association. Age, sex and number of MEG trials are included in each regression model, while IQ was added as an additional covariate for associations with neurodevelopmental symptoms.

**Figure 7.**
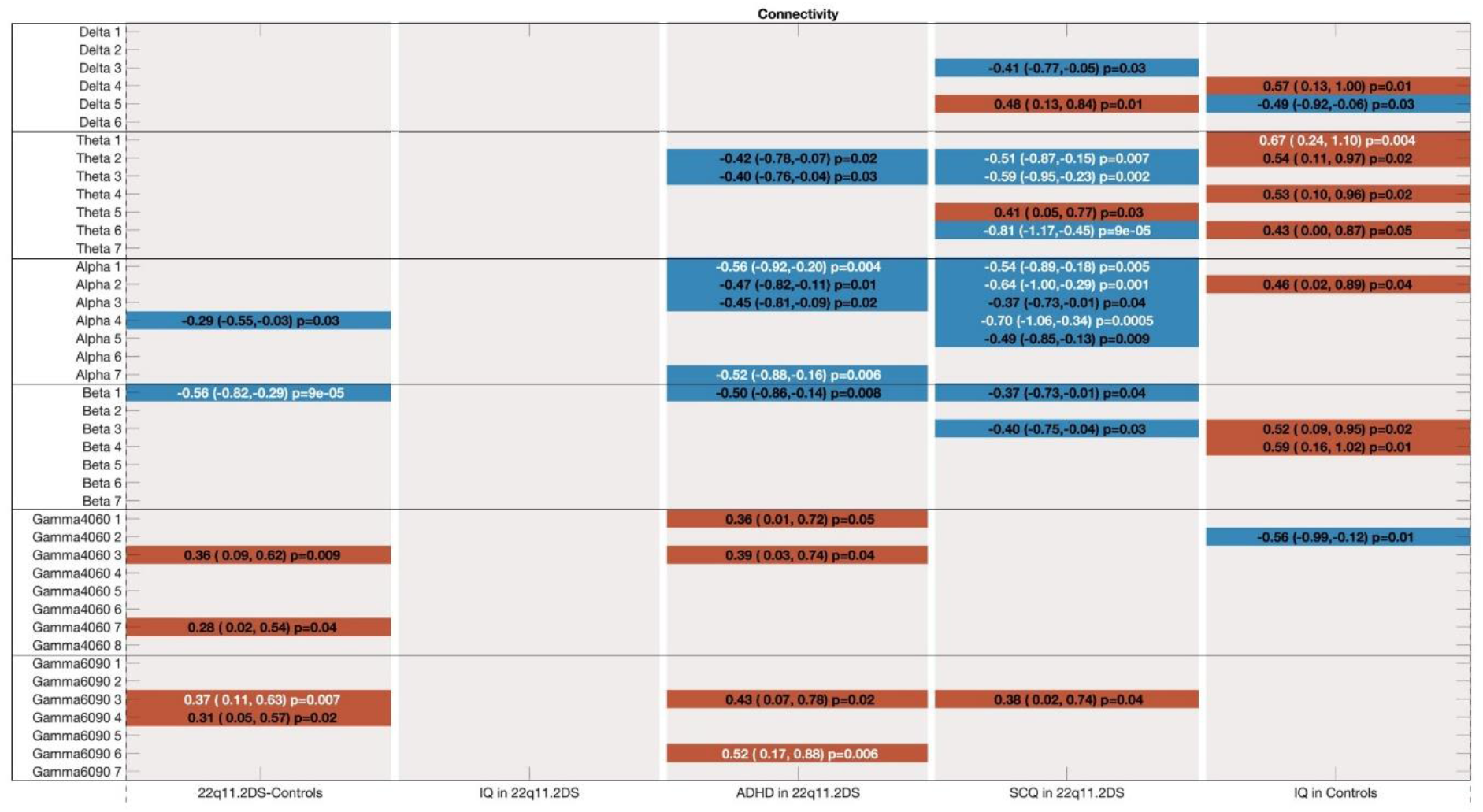
Significant associations between group status, IQ, neurodevelopmental symptoms and functional connectivity for each of the six frequency bands of interest. Components (p<0.05) showing positive associations are shown in red, while negative associations are shown in blue. Those associations surviving Bonferroni correction are highlighted in white text. Associations are shown by standardised beta values, confidence intervals and p values. Age and sex were included in each model as covariates, IQ was included as an additional covariate in models testing associations with SCQ and ADHD scores.

There were significant between-group differences in occipital beta and frontotemporal/frontoparietal high-gamma band functional connectivity. These differences are shown in figure 8. Beta band functional connectivity was lower in children with 22q11.2DS, while high gamma connectivity was higher in children with the CNV than in controls. As for the source activity comparisons, linear mixed modelling including sibling relatedness did not affect the significance of the results.

**Figure 8.**
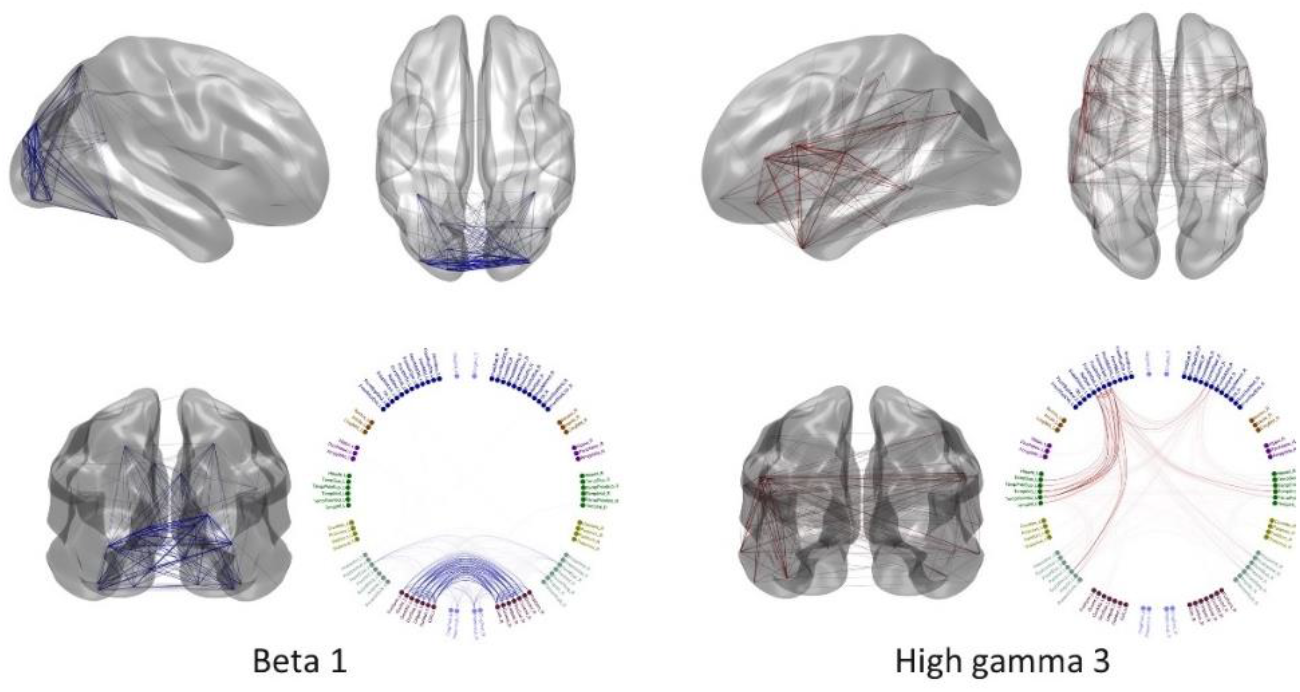
Locations of the two components showing significant between-group differences in functional connectivity between children with 22q11.2DS and controls, controlling for age, sex and number of trials. Blue indicates reduced connectivity and red indicates increased connectivity. Effect sizes: Beta 1 = -0.56 (p=0.000088), high gamma 3 = 0.37 (p=0.0069).

There were no significant associations between functional connectivity and IQ in children with 22q11.2DS. In controls, there was a positive association between IQ and theta band functional connectivity.

**Figure 9.**
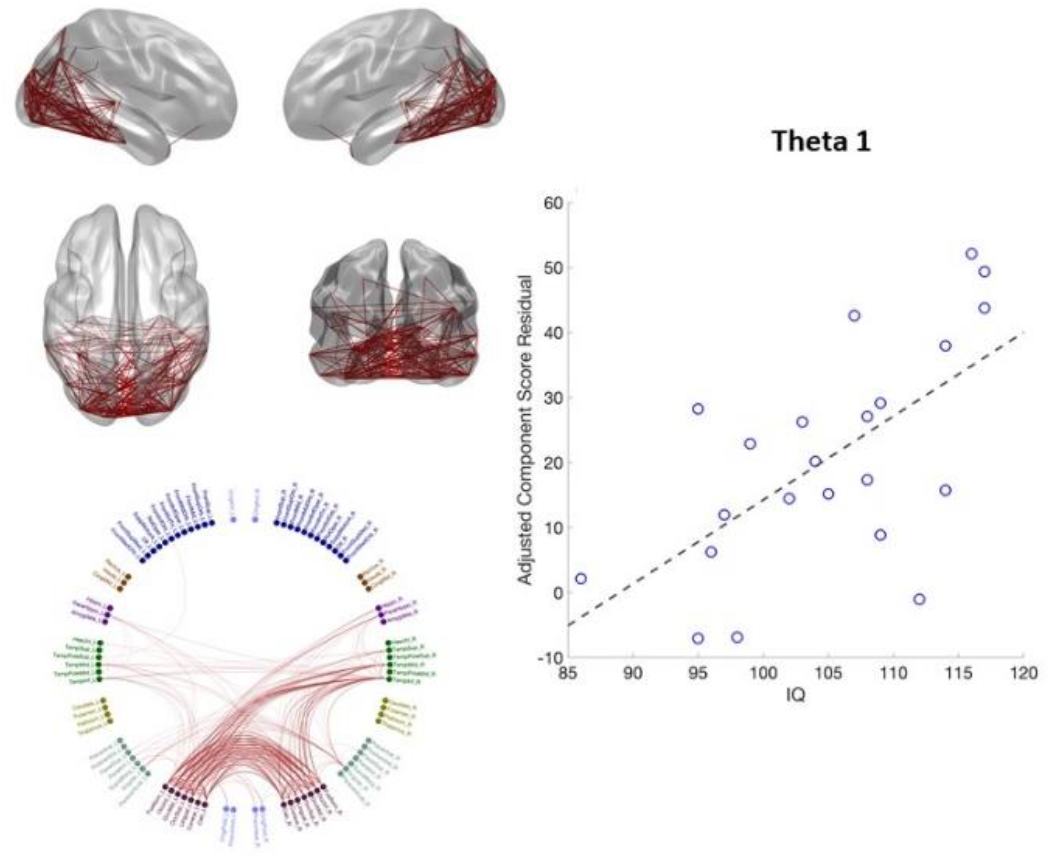
Relationship between IQ and theta functional connectivity in controls, controlling for age, sex and number of trials. Effect size = 0.66 (p=0.004). The red lines here indicate a positive association.

Social communication difficulties in children with 22q11.2DS were negatively associated with alpha band functional connectivity in components comprising nodes of the posterior default mode and sensorimotor networks.

**Figure 10.**
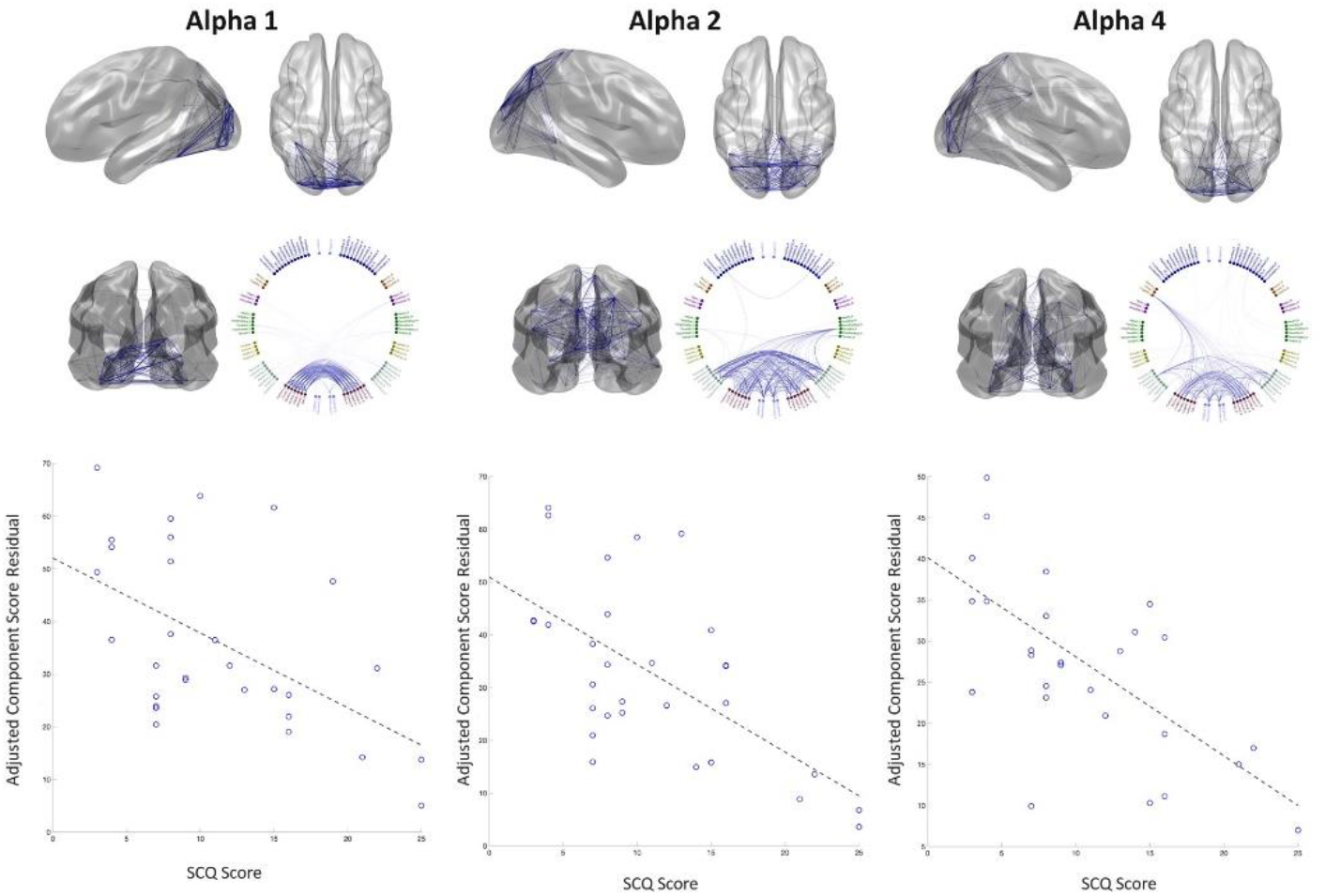
Relationship between alpha functional connectivity and SCQ scores, controlling for age, sex, IQ and number of trials. Effect sizes: Alpha 1 = -0.54 (p=0.005), alpha 2 = -0.64 (p=0.001), alpha 4 = -0.70 (p=0.0005). The blue lines here indicate a negative association.

ASD symptoms were also associated with alterations in theta band functional connectivity. Two of these networks (theta 2 and theta 6) are broadly similar to those seen in the alpha band, while theta 3 comprises frontal, temporal and occipital nodes.

**Figure 11.**
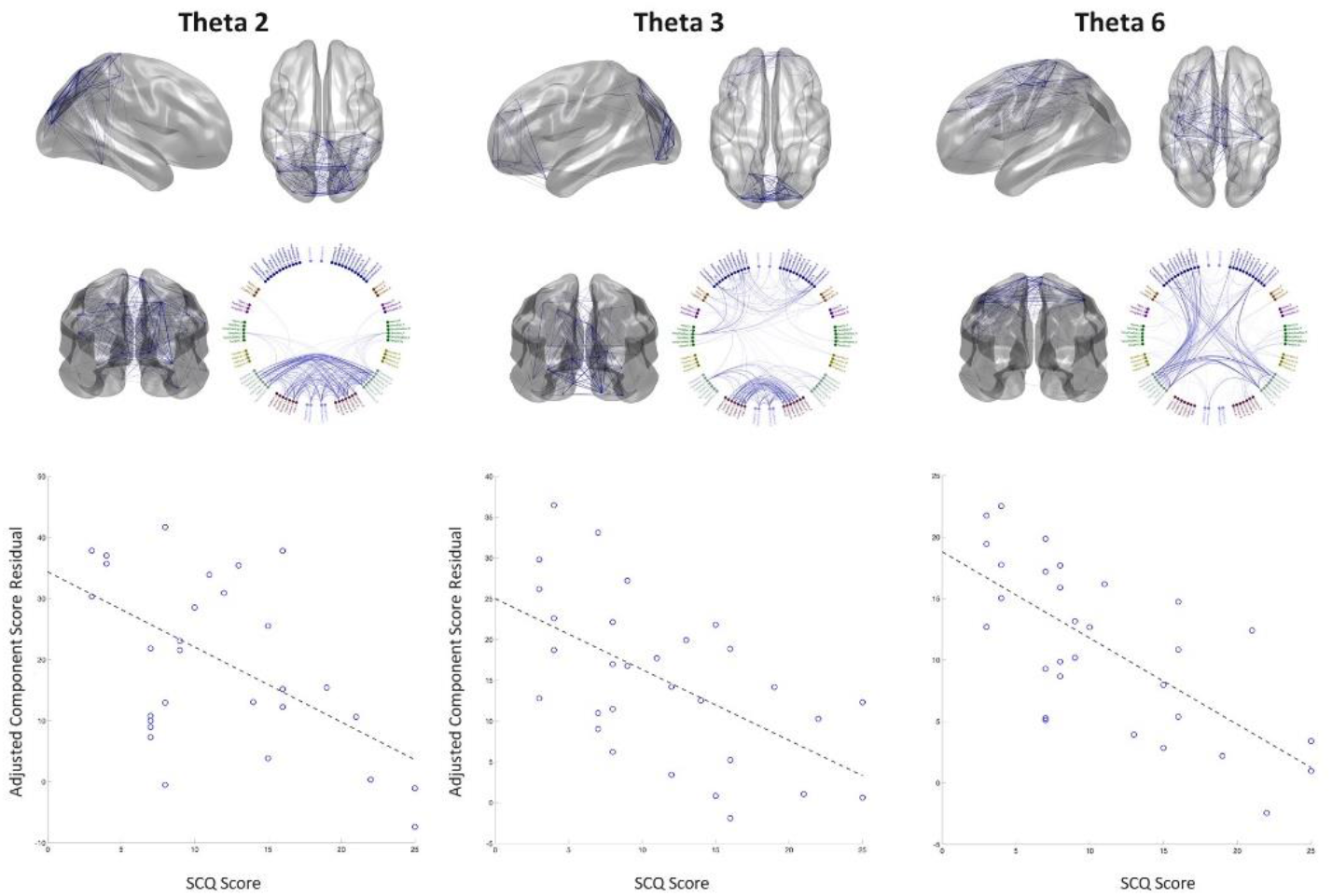
Relationship between theta band functional connectivity and SCQ scores, controlling for age, sex, IQ and number of trials. Theta 2 effect size = -0.51 (p=0.007), theta 3 effect size = -0.59 (p=0.002), theta 6 effect size = -0.81 (p=0.00009). The blue lines here indicate a negative association.

Finally, ADHD symptoms were associated with increased high gamma (60-90Hz) connectivity across a diffuse network including right frontotemporal, frontoparietal, temporoparietal and occipito-occipital connections. Alpha band functional connectivity was significantly reduced in two components, a posterior network consisting of occipito-occipital connections and an extended network involving multiple nodes across the brain.

**Figure 12.**
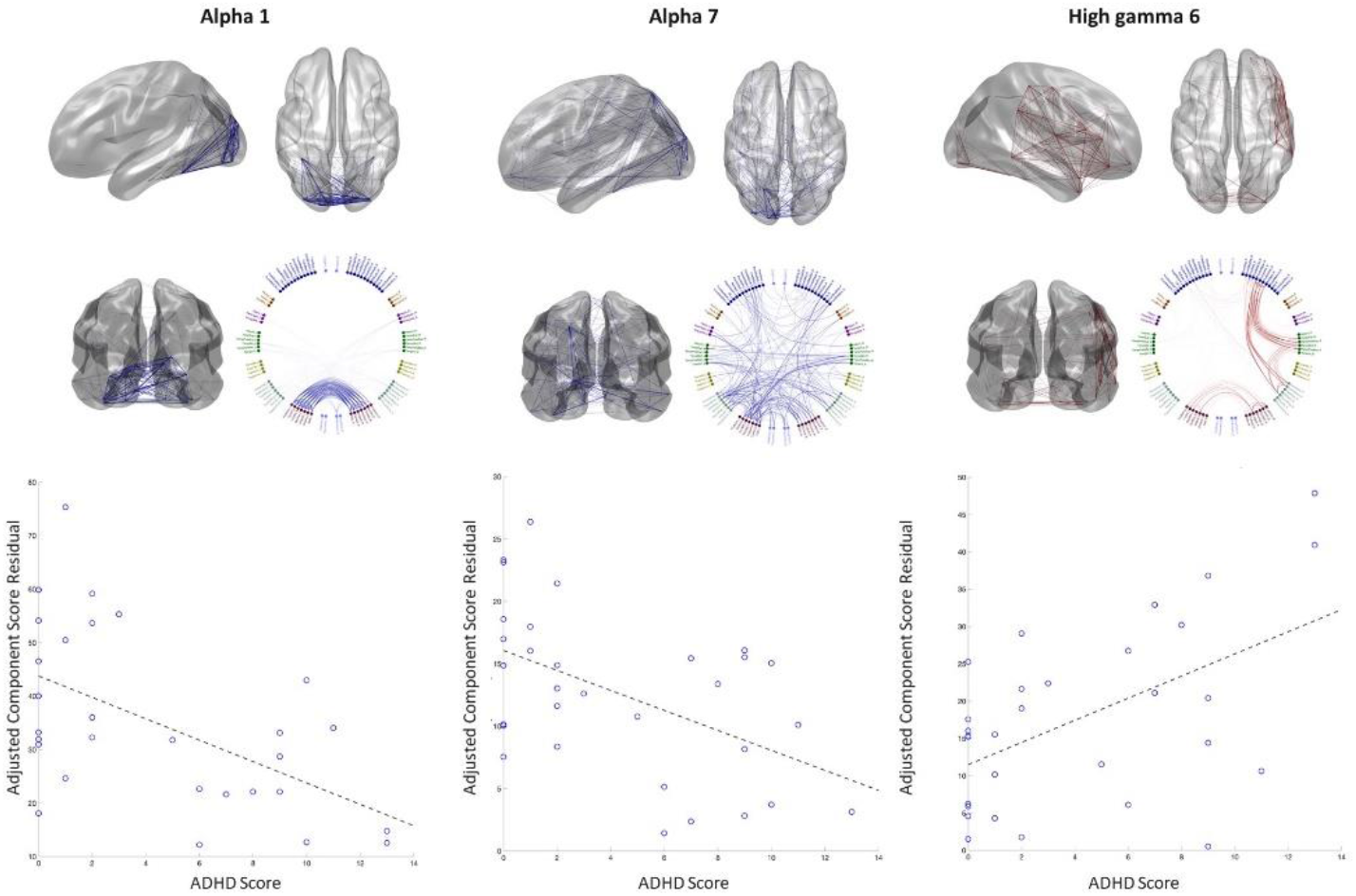
Relationship between functional connectivity and ADHD symptom scores, controlling for age, sex, IQ and number of MEG trials. Alpha 1 effect size = -0.56 (p=0.004), alpha 7 effect size = -0.52 (p=0.006), high gamma 6 effect size = 0.52 (p=0.006). Blue lines indicate a negative association and red a positive association.

## Discussion

22q11.2DS is a genetic syndrome associated with a variable and impairing neurodevelopmental phenotype. To our knowledge, this is the first study of resting-state neural oscillatory patterns in children with 22q11.2DS and the first study to explore the associations between oscillatory activity, functional connectivity and neurodevelopmental symptoms. Using robust between-group and exploratory within-group analysis strategies, we found atypical oscillatory patterns in children with 22q11.2DS, which were associated with cognitive impairments, social communication difficulties and ADHD symptoms. Of particular note, we found increased high frequency (gamma range) activity and functional connectivity in the frontal lobes of children with 22q11.2DS, which were associated with ASD and ADHD symptoms respectively. Conversely, occipital gamma band activity was reduced in 22q11.2DS and this was associated with ASD symptoms. Low frequency oscillatory activity and functional connectivity were also reduced in children with 22q11.2DS with reductions in low frequency activity being associated with cognitive difficulties and alterations functional connectivity being associated with neurodevelopmental symptoms. Our findings are in keeping with previous functional imaging studies in 22q11.2DS which report alterations in the frontal, default mode, sensorimotor and visual networks (23–27). These networks have also been implicated in idiopathic ASD (32,58–62) and ADHD ((63–67).

High frequency oscillations in the gamma range are thought to reflect local field potentials which arise as a result of the relative balance between excitation and inhibition in local cortical circuits (68). Cellular, animal and computational models have shown that these are largely driven by the action of parvalbumin-containing interneurons which synapse on excitatory pyramidal neurons (69–74). Excitatory-inhibitory imbalance has been proposed as a common neurobiological mechanism across the spectrum of neurodevelopmental disorders seen in 22q11.2DS (35,75–77). There are several mechanisms by which excitatory-inhibitory balance may be perturbed in 22q11.2DS. Firstly, haploinsufficency of *PRODH*, a gene in the 22q11.2 region, has been shown to affect GABA synthesis and gamma band activity in murine models (78). Secondly, these models have also found abnormal PV+ inhibitory interneuron migration in 22q11.2DS, a finding that is mirrored in postmortem studies of humans with 22q11.2DS (13–15,18,19). While there is little known evidence of regional brain differences in excitatory-inhibitory balance in 22q11.2DS, it is plausible that increased frontal and decreased occipital excitability could result in pleiotropic neurodevelopmental outcomes and psychosis risk. Indeed, increased frontal and decreased occipital gamma oscillations have been reported in patients with schizophrenia (79–81).

Low frequency oscillations are thought to facilitate long-range communication between brain regions (82). Computer simulations have further shown that low frequency amplitude envelope correlations seen in resting-state MEG studies may originate from spontaneous synchronisation mechanisms in the brain (83). Alterations in alpha, beta and theta band oscillations have been reported in ASD, ADHD and schizophrenia (67,77,84–89).

The functional connectivity changes we found in children are similar to those previously reported by our research group in adult neurodevelopmental CNV carriers. (41). While there were relatively few participants with 22q11.2DS in that study (n=14), analysis of the 22q11.2 group alone found similar connectivity reductions as the cross-CNV analysis but in addition, a number of nodes with hyperconnectivity were identified. 80% of participants in the adult study had a DSM-IV diagnosis (including 14% with schizophrenia) and 62% of the sample were taking medication. In the present study, while high rates of ASD and ADHD were found, none of the children had a psychotic disorder and all were free from psychotropic medication. Given that up to 30% of people with 22q11.2DS develop schizophrenia during their lifetime (11), it is highly likely that a proportion of the children recruited to this study will develop psychotic symptoms in the future. Following up these children over time may yield insights into neural markers of psychosis risk.

Strengths of this study include the relatively narrow age range recruited compared to previous electrophysiological studies of CNV carriers (10-17 years), clinical and cognitive phenotyping and robust MEG analysis strategies. However, there are also a number of limitations. Due to the relative rarity of 22q11.2DS, the sample size is modest. Therefore without replication, the findings should be interpreted cautiously, due to the risks of both type 1 and type 2 error (90). The physical health problems associated with 22q11.2DS meant that contraindications for MRI scanning were common and it was not possible to co-register MEG data to each participant’s own MRI scan. While every attempt was made to closely match the fiducial locations to those of another participant, this is not a perfect substitute for using participants’ own data and this may have affected source localization. The relatively short duration of the resting-state paradigm was chosen to balance data quantity and quality, however this will have resulted in lower SNR than longer paradigms (91). Unfortunately, a longer recording would not have been feasible for the majority of children taking part in this study. Furthermore, continuous head localization was not used during data acquisition so detailed analysis of head movement and correction for this was not possible. Children in both groups found it difficult to remain still during the recordings and therefore a pragmatic approach was taken to assessing data quality with higher upper limits of head motion being tolerated than in many studies involving adults.

In summary, the present study investigated resting-state neural oscillatory patterns in children with a genetic syndrome associated with neurodevelopmental disorders and cognitive impairment. We observed alterations in oscillatory activity/connectivity that were associated with the severity of cognitive difficulties, ASD and ADHD symptoms. These effects were seen at both high frequencies, thought to reflect local cortical processes, and at low frequencies, thought to reflect longer-range cortical communication. These results suggest a potential neural mechanism by which the 22q11.2 deletion could act to increase risk across the spectrum of neurodevelopmental disorders, involving anterior hyper- and posterior hypoconnectivity. This has implications for our understanding of 22q11.2DS, other neurodevelopmental CNVs and idiopathic neurodevelopmental disorders. In the future, longitudinal studies could shed light on how these oscillatory patterns may change during development and how they may relate to the emergence of psychotic symptoms.

## Data Availability

Data available on request, subject to data transfer agreements

## Acknowledgements

This work was supported by a Wellcome Trust ISSF Award and Clinical Research Training Fellowship to JD (102003/Z/13/Z), the Waterloo Foundation (code 918-1234) and the Baily Thomas Charitable Fund (2315/1) to MvdB and MJO, a Medical Research Council Centre Grant (No. MR/L010305/1) to MJO, an MRC MEG UK Partnership Grant to KDS (MR/K005464/1) and the National Centre for Mental Health. The data have previously been published on a preprint server.

The authors would like to thank Lisa Brindley, Suresh Muthukumaraswamy, Gavin Perry, Sonya Foley and Rachael Adams for their assistance with data collection, and Loes Koelewijn for her help with data pre-processing. They would also like to thank the charities and NHS clinics who helped to recruit participants and all of the families who participated in the study.

