## Supplementary material for "Alterations in resting-state activity and functional connectivity in children with 22q11.2 deletion syndrome"

**MEG analysis: Beamformer reconstruction**

Source reconstruction was performed for each participant band using a linearly-constrained minimum variance (LCMV) beamformer implemented in FieldTrip (version 20161011, www.fieldtriptoolbox.org (48)). Beamforming was performed in each of six distinct frequency bands using conventional definitions: delta (1-4 Hz), theta (4-8Hz), alpha (8-13Hz), beta (13-30Hz), low gamma (40-60Hz) and high gamma (60-90Hz). The source-model used for reconstruction was a 6mm isotropic grid, which was initially defined in MNI template space before being matched to each individual's MRI using Fieldtrip's *ft_prepare_sourcemodel* function. The head conductivity model used was the *localspheres* option in FieldTrip, which approximates the local curvature of the head underneath each channel (www.fieldtriptoolbox.org).

Beamformer weights were estimated for each location on the grid, via estimation of the covariance matrix over the entire resting-state recording period. After weights normalization, these weights were used to derive an estimated activity time series at each grid voxel and for each trial. These trial time series were concatenated to form a single time series for each grid voxel. After cleaning of any spike discontinuities using a temporal median filter, these time series were then taken forward for both activity and connectivity analyses.

**MEG analysis: Activity estimation**

For each of the reconstructed grid positions, a measure of activity was derived in each frequency band. This was done by first deriving the amplitude-envelope of the virtual-sensor time series using the absolute value of the analytic function transform of the raw time series (using Matlab's *hilbert* function). The resulting time series was down-sampled to a 1Hz, in order to match the connectivity analysis described below, and then converted to a single activity measure that summarizes how variable this envelope is over the entire resting-state run. To do this we calculated the coefficient-of-variation of the envelope, namely the temporal standard-deviation divided by the temporal mean. This normalized measure has the advantage of correcting for the known biases, introduced by the sensitivity of beamformer weights to variations in the signal to noise ratio (SNR) of the data (47,48). The end result is a 6mm isotropic activity map, for each participant and each frequency band.

**MEG analysis: Connectivity estimation**

Functional connectivity was computed using the amplitude-envelope correlation (AEC) metric. This metric has previously been shown to be both robust and repeatable (49). The analysis pipeline has previously been described (47). First, spatial down-sampling to the 90 regions of the Automated Anatomical Labelling (AAL90) atlas was performed (50). One grid source (virtual sensor) was chosen to represent each AAL90 region, based on the voxel having the largest temporal standard deviation across the resting-state experiment. The temporal activity of each of these 90 sources was then orthogonalized with respect to each other region in order to suppress any zero-time-lag correlation due to signal leakage (51). Next, the amplitude (Hilbert) envelopes of each AAL90 region were extracted using the absolute of the (complex) analytical signal derived by the *hilbert* function in MATLAB. These amplitude envelopes were then down-sampled to a temporal resolution of 1s in order to study connectivity mediated by slow amplitude envelope changes (52). A median spike removal filter was applied to smooth large deflections in the data before further analysis.

To obtain connectivity matrices, pairwise correlations were calculated between the 90 Hilbert envelopes, yielding 4005 unique correlations for each frequency band for each participant. Each of these correlation coefficients was then transformed to a variance-normalised Fisher z-statistic, using a procedure that estimates the temporal variance of the time series' null distribution for each region, using surrogates generated by randomisation. This made the correlations suitable for further statistical analysis and corrected for the varying length of the final time series for each participant.

**MEG analysis: Activity and connectivity component estimation**

At the end of the above analysis procedures, each participant had 6 activity maps (one for each frequency) and 6 connectivity matrices. Each activity map had 5061 voxels and each connectivity map has 4005 unique connection values. In order to reduce the dimensionality of these features before statistical analyses, we used a data-driven analysis of the principal components using non-negative matrix factorisation (Matlab:nnmf). This specific algorithm was chosen because the activity measure we have used is positive-only and the slow static amplitude-connectivity measure we have used is dominated by positive correlations. For interpretability, we also preferred each participant's loading on to each component to be a positive measure only. Recently, non-negative matrix factorisation has been successfully used to show cohort differences in a MEG study of schizophrenia (53) and in comparing structural and functional connectivity components in healthy individuals (54). One non-trivial issue in using NNMF is that it is difficult to decide how many components to reconstruct. Traditional stopping criteria, such as percentage variance explained, do not work well as the NNMF algorithm is able to significantly improve reconstruction accuracy by adding components that load on to small numbers of participants. We chose a heuristic approach in which we iteratively increased the number of components and tested what proportion of our cohort had non-zero values for each component. We required each component to be represented in at least 50% of our participants and, across all components, for the mean number of participants represented to be at least 70%. If, when we increased the number of components, either of these criteria was not met, we went back to the previous step. For each measure, we typically find that 5-15 components are identified to be the maximum number that meet these criteria. For each of the final components identified, we projected each individual's data on to these networks to get a single component 'strength' for each person. For each of the 12 metrics we have in each person (6 activity and 6 connectivity), we performed NNMF separately. As shown in figures 1,2 and 7, each participant's combined activity and connectivity profile, across all 6 bands, was effectively summarised by just 79 values. It is these values that were taken forward for statistical analysis.

**Statistical analyses of NNMF derived component scores**

Each of the component weightings described above was used in an analysis to determine whether their magnitude was predicted by a set of exploratory variables consisting of group-status (22q11DS or control), IQ and neurodevelopmental symptoms, using SCQ scores and CAPA-derived ADHD symptom counts to index the severity of ASD and ADHD symptoms respectively. Due to the differing IQ distributions in the two groups, associations with IQ were explored in each group separately. This analysis was done by a set of univariate robust general linear modelling tests, using Matlab's *fitlm* function. In each linear-model fit age, sex and number of MEG trials were included in the models as covariates. For linear models exploring the associations with ASD and ADHD symptoms, IQ was included as an additional covariate. In each test, we assessed the significance of the principal variable (Group, ASD in 22q11DS,

ADHD in 22q11DS, IQ in controls, IQ in 22q11DS) in explaining variance in the residuals after controlling for age and sex. With our relatively small participant numbers, outliers can have a strong effect on the quality of the model fit. We therefore used a form of robust fitting, using an iterative procedure, in which, after an initial fit, the residuals were assessed for outliers using Cooks' distance. We used a common combination of rules for outlier identification: i.e. if a participant's Cook's distance was greater than 3 times the cohort mean, or had an absolute value of greater than 0.5, the participant was excluded and the linear model was re-fit. This procedure was then repeated once more to generate the final model fit. Effect-sizes for the principal variable of interest were calculated using standardised-beta parameters and assessed for significance using p-values and 95% confidence intervals. Bonferroni correction was applied for the number of components tested within each type (activity/connectivity) and frequency-band i.e. correction was not applied across type and frequency band. To control for relatedness between sibling pairs in the sample, sensitivity analyses using linear mixed modelling were run in R Studio (Version 1.1.383 for Mac) for each of the significant components using the package lmerTest. In these analyses, component weighting was the variable of interest with group status, age, sex and number of MEG trials as fixed effects and family identification numbers as random effects.
